# Pancreatic cancer symptom trajectories from Danish registry data and free text in electronic health records

**DOI:** 10.1101/2023.02.13.23285861

**Authors:** Jessica Xin Hjaltelin, Sif Ingibergsdóttir Novitski, Isabella Friis Jørgensen, Julia Sidenius Johansen, Inna M Chen, Troels Siggaard, Siri Vulpius, Lars Juhl Jensen, Søren Brunak

## Abstract

Pancreatic cancer is one of the deadliest cancer types with poor treatment options. Better detection of early symptoms and relevant disease correlations could improve pancreatic cancer prognosis. In this retrospective study, we used symptom and disease codes (ICD-10) from the Danish National Patient Registry (NPR) encompassing 8.1 million patients from 1977 to 2018, of whom 22,727 were diagnosed with pancreatic cancer. To complement and compare these diagnosis codes with deeper clinical data, we used a text mining approach to extract symptoms from free text clinical notes in electronic health records (4,418 pancreatic cancer patients and 44,180 controls). We used both data sources to generate and compare symptom disease trajectories to uncover temporal patterns of symptoms prior to pancreatic cancer diagnosis for the same patients. We show that the text mining of the clinical notes was able to capture richer statistically significant symptom patterns, in particular general pain, abdominal pain, and liver-related conditions. We also detected haemorrhages (p-value = *4*.*80* · *10*^-*08*^) and headache (p-value = *2*.*12* ·*10*^-*06*^) to be linked as early symptoms of pancreatic cancer. Chaining symptoms together in trajectories identified patients with jaundice conditions having higher median survival (**>**90 days) compared to patients following trajectories that included haemorrhage, oedema or anaemia (≤90 days). Additionally, we discovered a group of cardiovascular patients that developed pancreatic cancer with a lower median survival (≤90 days). These results provide an overview of two types of pancreatic cancer symptom trajectories. The two approaches and data types complement each other to provide a fuller picture of the early risk factors for pancreatic cancer.

## Introduction

Pancreatic cancer has been predicted to become the second leading cause of cancer deaths, surpassing breast, colorectal, and prostate cancer(Rahib et al. 2021). It has few and generic symptoms resulting in late diagnosis(Kim and Ahuja 2015; Chari et al. 2015) and poor prognosis with a 5-year survival rate of 11%(“American Cancer Society” 2020). Hence, improved knowledge of symptoms and diseases occurring early is of high importance to treat this cancer type at a curable stage and provide better prognosis and guide screening programs for pancreatic cancer(Risch et al. 2015). If the cancer is detected at an early stage, where surgical removal of the tumor is possible, the survival rate increases to 42%(“American Cancer Society” 2020).

Symptoms of pancreatic cancer are often mistaken for signs of less severe illnesses and overlooked in clinical practice. Some of the most frequent symptoms linked to pancreatic cancer are weight loss, abdominal pain, and anorexia(Hidalgo 2010). Others include upper abdominal pain, cholestasis, nausea(Hidalgo 2010), and dark urine and thirst(Liao et al. 2021). Cholestatic symptoms are more commonly found when the tumor is located in the head of the pancreas(De La Cruz, Young, and Ruffin 2014)(Porta et al. 2005). New-onset diabetes has additionally been found to co-occur with pancreatic cancer when accompanied by weight loss(Yuan et al. 2020),(Hart et al. 2011),(Bruenderman and Martin 2015).

National or regional disease registries hold longitudinal data on disease development. The registries in the Nordic countries are of high quality and among the oldest covering treatment in one-payer health care systems(Laugesen et al. 2021). The National Danish Patient Registry (NPR) contains hospital diagnoses since 1977 and allows for large data-driven studies to detect temporal disease progression patterns relevant in the context of stratified medicine(Jensen et al. 2014; Siggaard et al. 2020). A recent example was the characterization of multimorbidity correlations across cancer types in 0.7 million patients(Hu et al. 2019). However, much of the deeper phenotypic patient information resides within the free text of the electronic health records (EHRs)(Soguero-Ruiz et al. 2016; Delespierre et al. 2017). A small-scale study using 4,080 mixed types of cancers attempted to build more general “event trajectories” using a text mining and a pooled analysis-approach (Jensen et al. 2017). A prospective study investigating initial symptoms and diagnostic interval (time from onset to diagnosis) for known pancreatic cancer symptoms found no difference between pancreatic cancer and patients suspected of having pancreatic cancer(Walter et al. 2016). It is also suggested that symptoms appear sporadically, adding to the complex nature of the disease manifestation(Evans et al. 2014). Other studies used primary care EHRs to detect pancreatic cancer symptoms and found jaundice(Stapley et al. 2012), back pain, lethargy, and new-onset diabetes to be linked to pancreatic cancer(Keane et al. 2014). These studies detected pancreatic cancer symptoms using a single-disease approach, not considering the temporal ordering of symptoms or diseases.

In this paper, we present a large-scale study to investigate pancreatic cancer symptoms longitudinally. We cover all symptoms included in the International Classification of Disease (ICD-10) terminology symptom chapter 18. Additionally, we also include in the text mining vocabulary other known or suggested pancreatic cancer symptoms. We generate and compare disease and symptom trajectories using registry data and clinical notes in EHRs to characterize the temporal ordering of symptoms across data sources.

## Results

### Extracting patient-level data from the Danish National Patient Registry and free text electronic health records

The Danish National Patient Registry (NPR) data spans the period 1977 to 2018, while the electronic health records (EHRs) used here are from the 2006 to 2016 time-interval. The NPR includes 8,110,702 patients where 22,727 patients are diagnosed with pancreatic cancer (**Table 1**). A subset of 4,418 of these pancreatic cancer patients is included in the EHRs. Almost as many females as males are identified with pancreatic cancer both in NPR and the EHRs (**Table 1**).

**Table 1.** Data set and patient characteristics.

| Data set | The National health Registry (NPR) | The free text clinical notes |
| --- | --- | --- |
| Year span | 1977-2018 | 2006-2016 |
| N pancreatic pancer patients | 22,727 | 4,418 |
| N controls | 8.1M | 44,180 |
| Female | 11,326 (49.8%) | 2,138 (48.4%) |
| Male | 11,401 (50.2%) | 2,280 (51.6%) |
| Mean age at diagnosis (female/male) | 73/70 | 72/70 |
| Counts age at diagnosis |  |  |
| <40 | 232 | 33 |
| 40-50 | 914 | 161 |
| 50-60 | 2,998 | 520 |
| 60-70 | 6,165 | 1,355 |
| 70-80 | 7,564 | 1,492 |
| >80 | 4,854 | 857 |

We limited the pancreatic cancer patient symptom history in the NPR and clinical notes to five years prior to the diagnosis. The text-based approach was able to identify 132 unique symptoms in the clinical notes, not registered in NPR (**Fig. 1A**). The most frequent symptoms exclusively found in the clinical notes were abnormal blood pressure, anaemias, and conditions related to the intestine (**Fig. 1B**). Within abnormal blood pressure, 1,656 patients had hypertension and 75 patients had low blood pressure. In addition, emotional states like unhappiness and worries were also found (**Fig. 1B**). NPR contained 122 ICD-10 symptoms not found by text mining (**Fig. 1A**). Frequent symptoms exclusively identified in the NPR data comprise abdominal pain, dyspnea, pain localized to the upper abdomen, and abnormal findings on medical images (**Fig. 1C**). Despite the clinical notes data set having much fewer pancreatic cancer patients (4,418 versus 22,727 in NPR), we found a significantly higher number of symptoms in these. In NPR and free text clinical notes, 185 symptoms were identified that occurred in both sources (**Fig. 1A**). Of these 185 symptoms, the top 10 most frequent symptoms from the free text clinical notes and the NPR were compared (**Fig. 1D**). Both sources agreed on six symptoms as being among the top 10 most frequent symptoms for pancreatic cancer patients. These six symptoms were jaundice, abnormalities of breathing, dizziness and giddiness, abdominal and pelvic pain, symptoms and signs concerning food and fluid intake, and pain. From the hierarchical structure of the ICD-10 chapters, different levels of coding detail can be retrieved. In the symptom group “symptom and signs concerning food and fluid intake”, the majority of patients represent the subgroups “abnormal weight loss” (R63.4) and anorexia (R63.0). Nausea, vomiting, anorexia, and haemorrhage were frequent symptoms in the free-text clinical notes but have low occurrences in NPR. On the contrary, acute abdomen was frequent in NPR but barely found in the clinical notes (**Fig. 1D**).

**Fig. 1.**
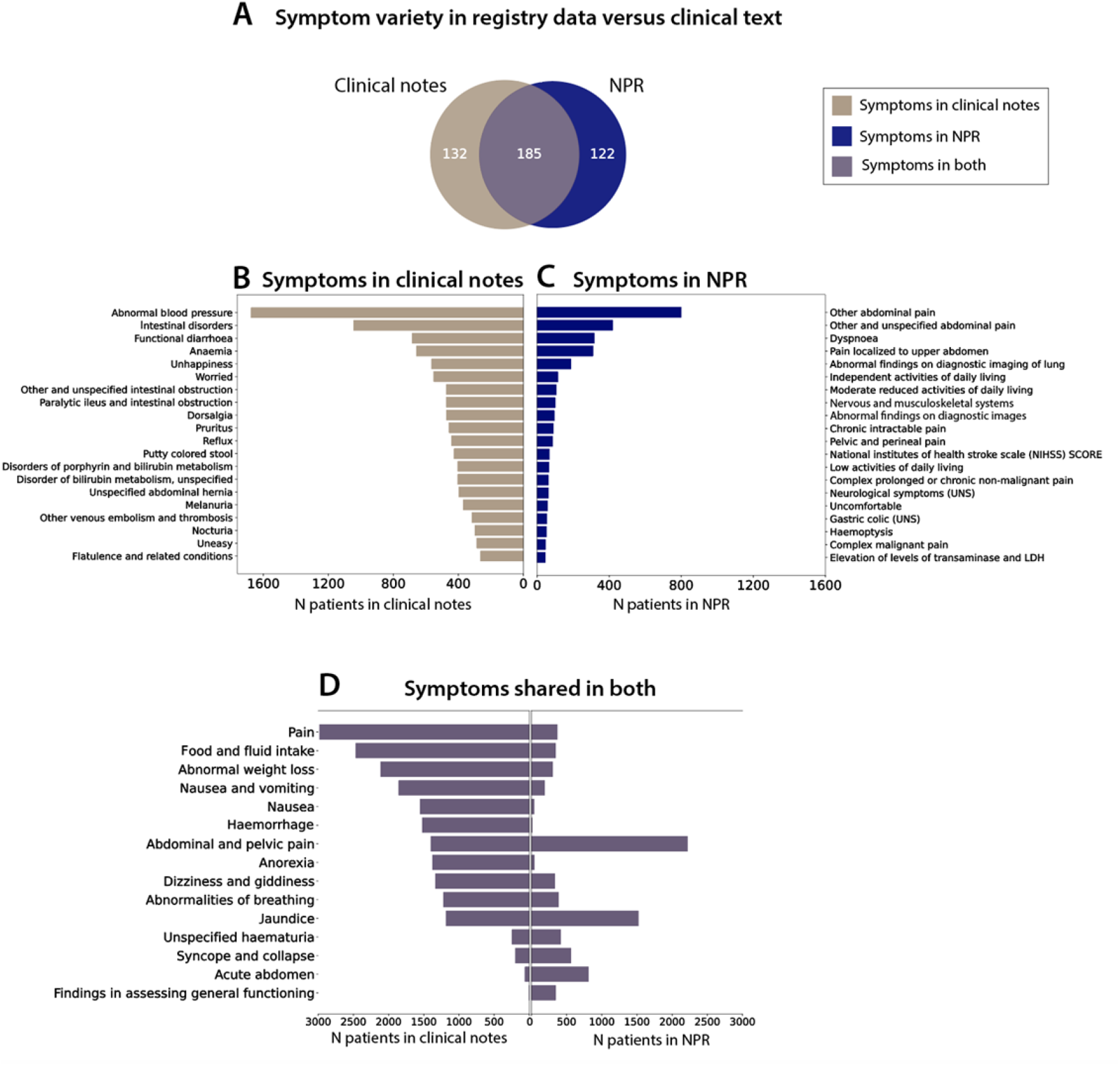
Comparison of pancreatic cancer symptoms in the Danish National Patient Registry (NPR) and electronic health records (EHRs). (**A**) Symptoms previous to the pancreatic cancer diagnosis identified in NPR (N_NPR_=122), by text mining of clinical notes from EHRs (N_notes_=132) or in both data sources (N_both_=185). (**B**) The top 20 most frequent symptoms that are only found in the clinical notes. (**C**) The top 20 most frequent symptoms only found in NPR. (**D**) The top 15 most frequent symptoms from the clinical notes and NPR, from the list of 185 overlapping symptoms. Some symptom names have been shortened for overview (see Supplementary Table S1).

### Text mining validation

In total, 4,141 out of 4,418 (94%) of the pancreatic cancer patients in clinical notes have at least one symptom identified by text mining. A control group of 44,180 patients was generated by matching age and sex. From these, 38,503 patients (87%) had a match for at least one symptom. The performance of the text mining was validated using a test corpus comprising a random extraction of 200 clinical notes from 200 different patients. In these notes, a total of 807 symptoms were manually annotated and the text mining method was able to identify 675 correctly and 132 symptoms were not found. This yielded a sensitivity score of 83.4%. A specificity score of 99.8% was obtained since the majority of clinical notes comprise non-symptom words describing the patient’s contact with the hospital. Symptoms incorrectly matched to the dictionary constitute a total of 53 words.

### Pancreatic cancer trajectories from NPR

Longitudinal disease trajectories were generated for the pancreatic cancer patients, where significant directional diagnosis pairs were joined to represent patients that traverse a complete disease path. The width of the trajectories illustrates the size of a patient group that moves through a particular path and a patient can follow multiple paths. The ICD-10 disease codes from NPR were used to generate disease trajectories (**Fig. 2A, Supplementary Table S2**). ICD-10 chapters, which are associated with pancreatic cancer, are abdominal diseases, cardiovascular diseases, diseases relating to the ears and mastoid process, endocrine, nutritional and metabolic diseases, and the symptom chapter. Most of the codes from the symptoms chapter appear after the diagnosis of pancreatic cancer, except for abdominal and pelvic pain (**Fig. 2A**). This symptom is part of six different trajectories comprising 1,363 patients. One of the trajectory groups is formed by cardiovascular diseases (1,720 patients) including angina pectoris and acute myocardial infarction that traverse to angina pectoris, chronic ischemic heart disease, type 2 diabetes or heart failure and afterwards into malignant neoplasm of the pancreas. The patients following these trajectories have the shortest survival (median survival ≤90 days) compared to the patients following the other trajectories. Other disease trajectories that show short survival are the cataract trajectory and the gonarthrosis-hypertension trajectory, comprising 321 and 332 patients, respectively. The rest of the trajectories has median survival above >90 days and includes for example type 2 diabetes (780 patients) or abdominal pain (1,363 patients) before pancreatic cancer. Survival in months for the patients following a disease trajectory is shown in **Supplementary Fig. S1B**.

**Fig. 2.**
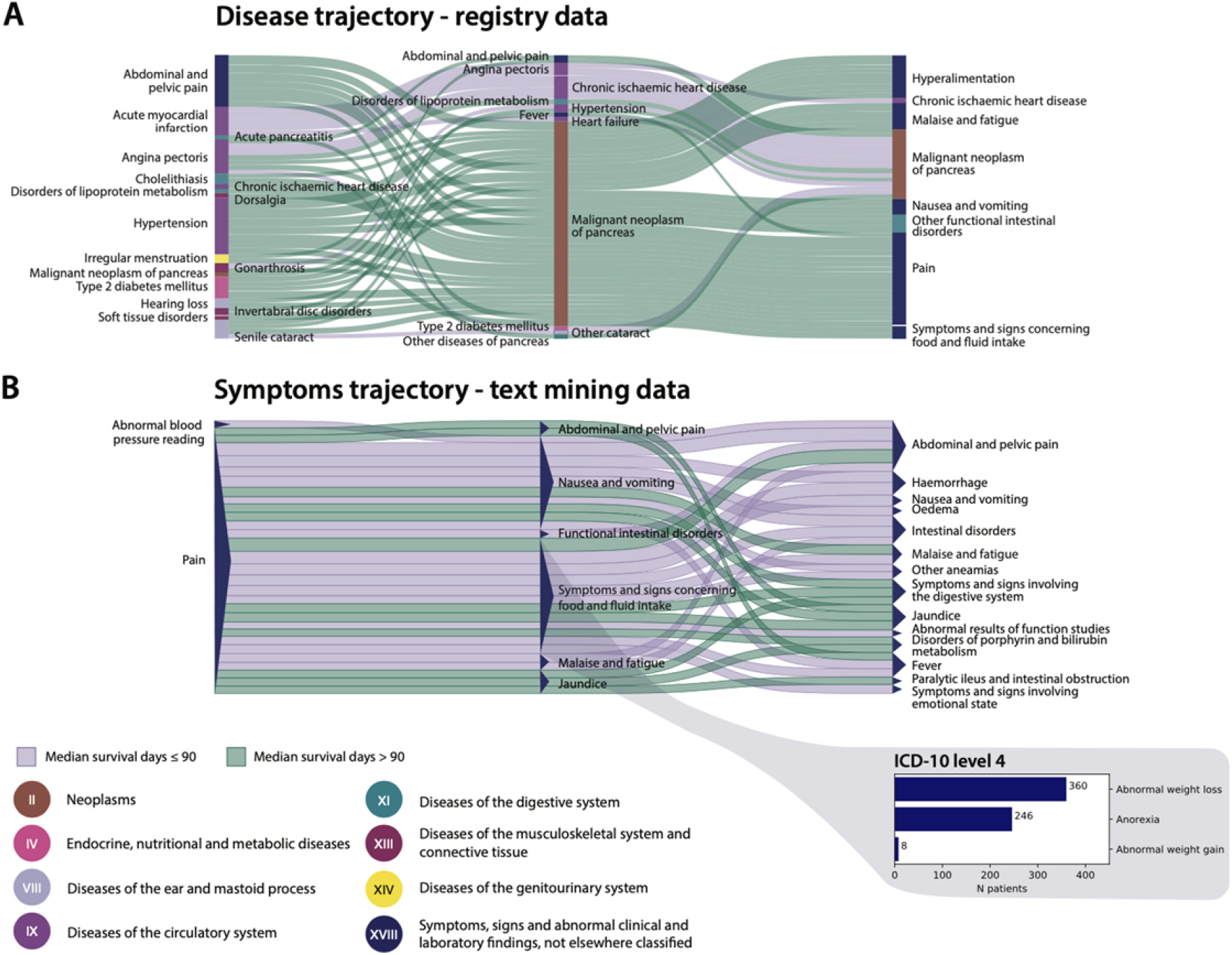
Disease and symptoms trajectories before and after pancreatic cancer diagnosis. **(A)** The registry disease trajectories consist of significant disease pairs with a Relative Risk (RR) > 1 (Supplementary Table S1). Each trajectory has a minimum of 300 patients. **(B)** Symptom trajectories from clinical notes consisting of significant disease pairs with RR > 1 (Supplementary Table S3). Each trajectory has a minimum of 100 patients. The width of the trajectories indicates visually the actual number of patients. The purple-colored trajectories represent patient groups with median survival ≤90 days. The green-colored trajectories represent patient groups with median survival >90. Disease and symptoms in the trajectories are colored by the ICD-10 chapters. Some symptoms names have been shortened for overview.

### Pancreatic cancer trajectories from clinical notes

The symptom trajectories were generated for the pancreatic cancer patients with available clinical notes 5-years prior to the diagnosis (**Fig. 2B**). All trajectories shown are significant in terms of direction with at least 100 patients following the complete path of three symptoms. A patient can follow several symptom paths. The majority of the trajectories begin with pain including 1,085 patients. From the pain symptom, a large group traverse into symptoms and signs concerning food and fluid intake (N=614). Of these 614 patients, 360 have abnormal weight loss, 246 have anorexia, and eight have abnormal weight gain. Several trajectories were also identified with pain that traverse into symptoms of nausea and vomiting or jaundice. At the end of the three symptom trajectories, 376 patients end up with abdominal pain, 272 patients with haemorrhage, 261 with functional intestinal disorders, and 222 with jaundice. All the patients following trajectories that end up with haemorrhage, oedema, and anemia have short survival (median survival ≤90 days) opposed to for example disorders of porphyrin and bilirubin metabolism and jaundice (median survival >90 days). See **Supplementary Table S3** for trajectory details. The survival of the patients following the symptom trajectories are presented in **Supplementary Fig. S1A**.

### Temporality of symptoms from clinical notes

The distribution of symptoms registered over time can be seen for the 20 most frequent symptoms that occur significantly more often in the pancreatic cancer patients opposed to the control group (**Fig. 3**). The distribution is over a five-year period previous to the pancreatic cancer diagnosis. If a symptom is registered multiple times in one hospital encounter it is included once. Otherwise, all occurrences of a symptom during the five-year period are included for a patient. Some of the most frequent symptoms identified are symptoms related to pain, weight loss, and jaundice. Additionally, haemorrhage (p-value = *4*.*80* ·*10*^-*08*^) and headache (p-value = *2*.*12* ·*10*^-*06*^) were frequent and significant among pancreatic cancer patients. **Supplementary Table S4** shows the number of patients and all p-values relating to these symptoms.

**Fig. 3.**
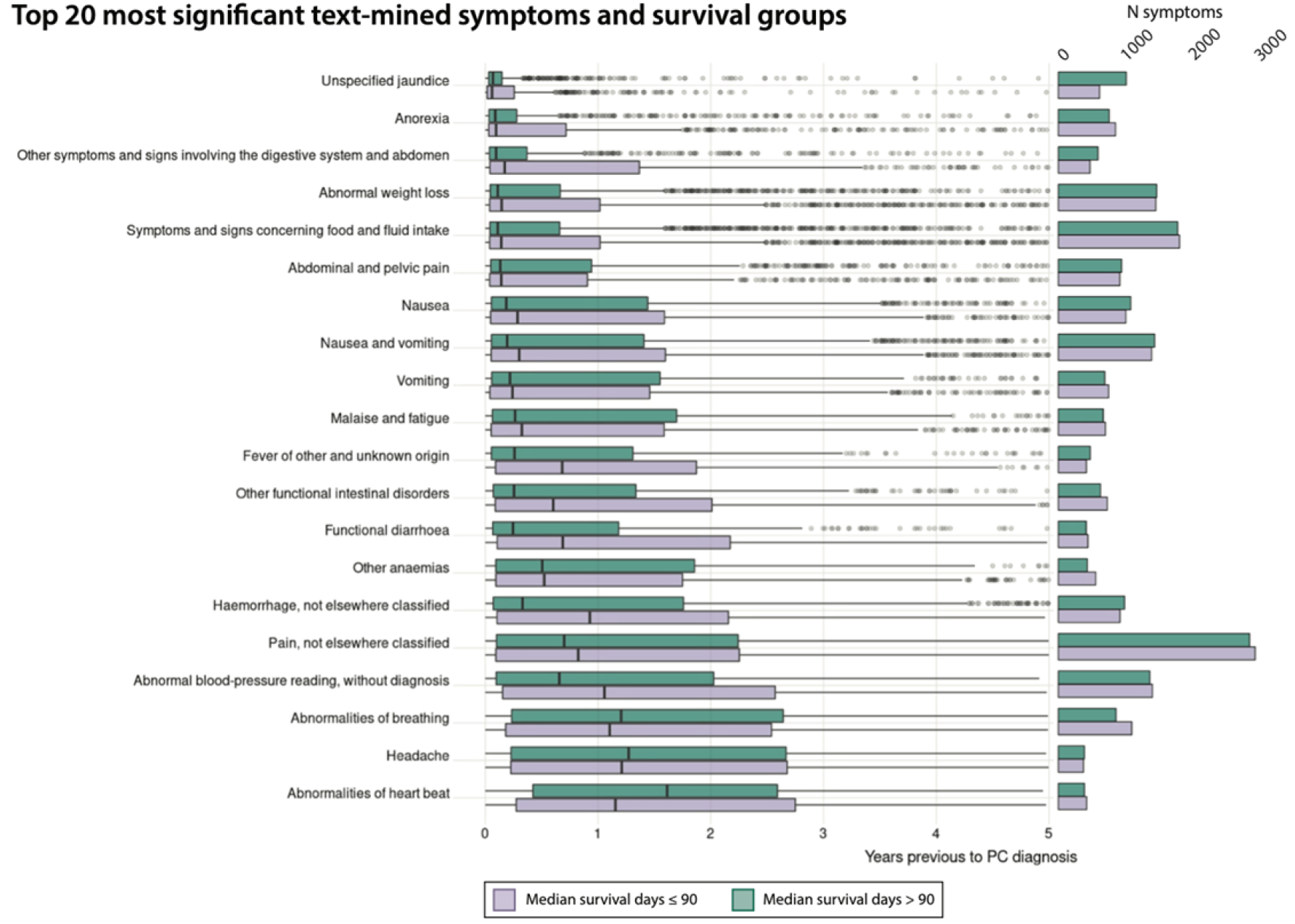
Top 20 most frequent text mined symptoms from the clinical notes five years previous to pancreatic cancer diagnosis. The top 20 most common and significant (P<0.05) symptoms in the text mined clinical notes are shown with survival information and time to pancreatic cancer diagnosis (Supplementary Table S4 and S5). The symptoms are extracted over a five-year period up to the time of pancreatic cancer (PC) diagnosis. If a symptom is noted more than once in one hospital encounter, the symptom is counted once. The purple bars indicate patients with survival ≤90 days and the green bars indicate patients with a survival >90 days. Symptom names may have been shortened for overview (see Supplementary Table S1).

Symptoms such as jaundice, anorexia, weight loss, and abdominal conditions were observed only closer to the pancreatic cancer diagnosis and the median of these appear within 6 months before pancreatic cancer diagnosis. The more general symptoms such as pain, abnormalities of breathing, abnormal blood pressure, abnormalities of heart beat, and headache have a median higher than 6 months before diagnosis. Most of the symptoms can be identified earlier in the clinical notes for patients with short survival (≤90 days) opposed to patients with longer survival (> 90 days).

## Discussion

This study uncovered statistically significant disease and symptom trajectories prior to pancreatic cancer that may be further assessed as early risk factors for pancreatic cancer for screening purposes. We complemented pancreatic cancer symptoms from hospital diagnosis codes with symptoms extracted by a text mining approach using free text in EHRs. From this, we discovered that symptoms were more abundant in the EHRs opposed to the NPR. The high variation in symptom frequency between the two sources was astonishing since the clinical notes were only available on a subset of the pancreatic cancer patients (4,418 out of 22,727). For example, more than 2,000 patients out of 4,418 had weight loss according to the free text clinical notes but in the registry, it was less than 500 patients out of 22,727. On the contrary, there were symptoms identified in the NPR that were not identified in the clinical notes which could be explained by the higher number of cases over a larger period of time in the NPR data set.

The most common pancreatic cancer symptoms such as abdominal pain and weight loss (Porta et al. 2005) as well as jaundice(Walter et al. 2016) were detected in both the registry and the text mining approach. Another study found that 21% of pancreatic cancer patients had dyspnoea(Krech and Walsh 1991), which agrees with our findings in relation to abnormalities of breathing. However, we discovered that haemorrhage and headache were frequent and significant in this cohort which is usually not described as a typical symptom of pancreatic cancer. In the clinical notes, more than a third of the patients had haemorrhages and the incidence is reported to be only 3%-12% for advanced cancers(Harris and Noble 2009). Additionally, gastrointestinal bleeding has been found to occur rarely among pancreatic cancer patients(Lee et al. 1994; Richardson and Baldeo 2016). Headache has been found to occur in 11% of advanced cancer patients which fits roughly with our results(Walsh, Donnelly, and Rybicki 2000), but is usually not described in relation to pancreatic cancer. Treatments or other comorbidities are entities that could be considered in relation to these findings to check if it is caused by the cancer, external source or by other confounders.

We found that for patients with short survival (≤90 days), symptoms could be tracked further back in the clinical notes, where patients with longer survival showed symptoms appearing closer to diagnosis. This could indicate that the pancreatic cancer had metastasized further in the patients with short survival. Short survival is commonly defined in oncology studies as death within 90 days(Sgouros and Maraveyas 2008). Using this threshold could be a strict choice to distinguish patient groups and it is important to interpret survival of each patient group exactly.

Tables S2 and S3 provide an overview of all the trajectories identified in our study including how many patients follow them and the median survival of each trajectory group. Symptom trajectories with a shorter median survival (≤90 days) included anaemia, haemorrhages, and oedema which is related to advanced cancers(Caro et al. 2001), (Pereira and Phan 2004), (Tai et al. 2016). Jaundice was coupled to higher survival (>90 days), since in the case of pancreatic cancer, it contributes as a clearer cancer symptom compared to the other more generic pancreatic cancer symptoms(Strasberg et al. 2014). Another study found the diagnostic interval to be shorter for jaundice compared to for example weight loss for pancreatic cancer patients(Walter et al. 2016), (Gobbi et al. 2013). It could therefore indicate a faster diagnosis but not necessarily a longer survival. We additionally identified patients with heart diseases and a lower survival that developed pancreatic cancer and it has been suggested that thromboembolic events could play a role in pancreatic cancer(Bergqvist et al. 2006) (Bertero et al. 2018). Cardiovascular diseases have been indicated as a risk factor for certain cancers(Lau et al. 2021), and contrariwise it is discussed if cancer should be included in cardiovascular risk prediction tools(Blaes and Shenoy 2019). Though the aim in our study was to identify patient groups that follow specific symptom trajectories there seem to be no prominent groups that stand out. All patients begin with general symptoms such as pain or abnormal blood pressure and afterwards more cancer specific symptoms such as weight loss, nausea, and jaundice appear.

One limitation of this study is that our data is strictly hospital contacts, which may result in a set of identified symptoms and diagnoses occurring not at the earliest disease stage. By including data from general practitioners, future studies could potentially identify even earlier symptom patterns prior to pancreatic cancer. Causal patterns reflected from trajectories can be difficult to interpret for example if an event serves as an actual cause or is the result of confounders(Jensen et al. 2014). Confounders could for example be medication or other risk factors that are not accounted for in the analyses. In this type of trajectory study, we can only assess the association between diseases, but actual causal relationships will need to be validated in future studies. Furthermore, the available period for the clinical notes (2006-2016) was shorter compared to the NPR (1977-2018). A direct comparison between the two data sources can be challenging, since registry terms are not necessarily written as in the free text clinical notes. One example is acute abdomen, which describes a condition with severe abdominal pain that demands immediate medical attention. A clinical note might contain text that the patient was admitted with severe stomach pain but register it as the ICD-10 code “R10.0 - Acute abdomen” in NPR which essentially would mean the same. Also, information bias may exist in the coding procedure within NPR and it might not always be trivial to code the correct symptom or diagnosis(Schmidt et al. 2015), (Lynge, Sandegaard, and Rebolj 2011). From our study, we show that the inclusion of symptoms text mined from clinical notes largely complemented ICD-coded symptoms from the patient registry.

This study showed that deep phenotypical information stored in registry data and free text EHRs can be useful for detecting temporal patterns. The sequence of events that leads up till the pancreatic cancer diagnosis supports that symptoms may appear in a messy and complex order. Patients start experiencing general and unspecific symptoms, which then become more specific and severe as the cancer advances. The methodology presented here can be used for external data sets and may, when validated, serve as a clinical tool to stratify patient groups so the correct option of patient care can be offered at a personalized level.

## Methods

### Study design

The health data used was from the Danish NPR and from clinical notes. For the generation of the trajectories, we used disease codes from the hierarchical International Classification of Diseases (ICD) system. ICD version 10 codes at level 3 were used for this study. The case cohort of pancreatic cancer patients were defined based on the pancreatic cancer code C25. If patients were registered with another cancer type before pancreatic cancer, they were not included as cases.

### Patient cohorts

From the Danish NPR, all hospital encounters in Denmark during the period 1977 to 2018 were used in our analyses, comprising 235,648,697 encounters for 8,172,661 patients. The ICD-10 Chapter 18 (Symptoms, signs and abnormal clinical and laboratory findings, not elsewhere classified) was used as vocabularies for extraction of symptoms from both NPR and the clinical notes. The EHR data comprising clinical notes included 1,906,769 patients from the Capital Region of Denmark and Region Zealand from the period of 2006 to 2016 with a total of 7,831,920 unique clinical notes. A clinical note contains information regarding a hospital encounter for a patient such as the reason for admission, findings, symptoms, operations, and treatments. Only patients with at least one clinical note within a five-year time interval before diagnosis were included. A control group was sampled for the patients having clinical notes using the same age and sex distribution with 10 control patients per pancreatic cancer patient and the same filtering criteria. The registries could be linked through the Central Personal Register (CPR) number, which all Danish citizens possess.

### Preprocessing of patient cohorts

Status codes 01 and 90 have been included in the analysis, excluding the rest of the status codes to make sure only active residents in Denmark (01) and inactive dead (90) are a part of the cohort. Furthermore, diagnosis types H and M (referral and temporary diagnosis) were excluded from the data for the extraction of disease and symptom frequencies and for the construction of trajectories. This was to ensure that the analysis is based on the main biologically relevant diagnoses in NPR.

### Text mining clinical notes

A dictionary was constructed by including all symptoms from the ICD-10 symptom chapter 18. Known pancreatic cancer symptoms that were not already a part of chapter 18, were added to the vocabulary. The final symptom dictionary consisted of 691 symptoms. The symptoms were initially written in their singular forms and afterwards suffixes were added to ensure different variants of a symptom such as plural forms. Other word endings like “condition” or “symptom” were also added since these are sometimes put behind a symptom in Danish (**Fig. 4**). Afterwards, the extended dictionary and the clinical notes from the patients and the control group were tokenized. The unique tokens from each were then compared to extract spelling errors on words longer or equal to five characters. In order to extract words with spelling errors, the tokens from the dictionary were fuzzy matched against the tokens from the clinical notes using the Python package fuzzysearch(Einat 2020).

**Fig. 4.**
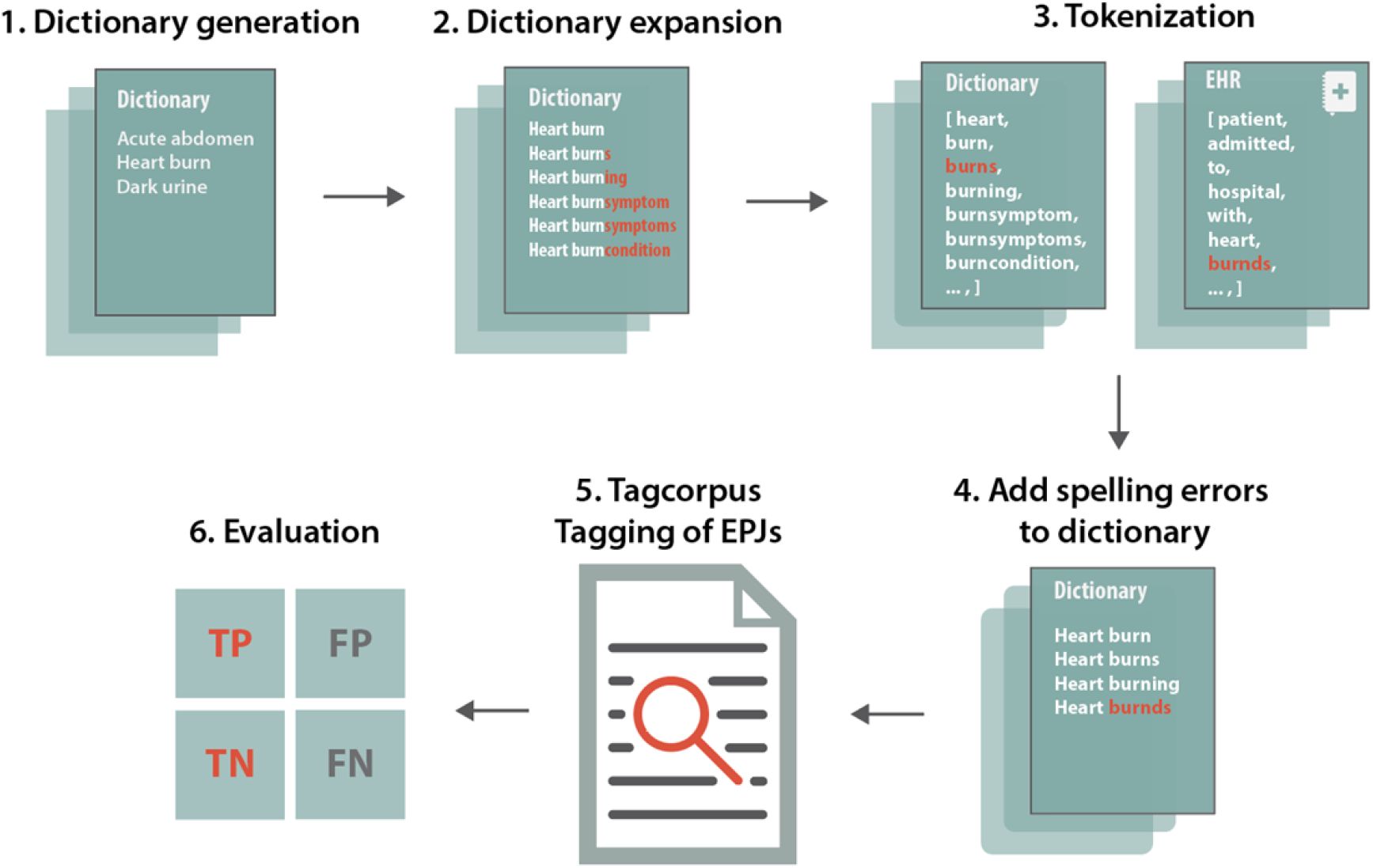
Text mining pipeline. A dictionary was generated with symptoms and expanded with word endings to capture multiple forms of the same symptom. Afterwards, the dictionary and the corpus (clinical notes) were tokenized in order to extract spelling errors. The spelling errors were then added to the dictionary. Finally, the program tagcorpus (Pafilis et al. 2013) was used to tag the symptoms in the corpus. The text mining performance was then evaluated.

The metric to measure similarities was Levenshtein’s distance. In this case, a maximum value of distance between two words was set to 1 to allow for either one substitution, one deletion or one addition of a character to the word. When the fuzzy matching was complete, all outcomes were further assessed and rechecked so that wrong matches could be removed.

After the extraction of variations for the synonyms, the program Tagcorpus(Pafilis et al. 2013) was used to tag the corpus (the clinical notes) for the pancreatic cancer patients and the control patients. It is a fast program written in C++, which for example is able to process thousands of PubMed abstracts per second((Pafilis and Jensen 2016), (L. J. Jensen 2016),(Pafilis et al. 2013)). A post-processing step was applied that removed sentences with negations and mentioning of other persons than the patient(Eriksson et al. 2014). This ensures that whenever there is a negation word in a sentence the word will not be tagged. For example, not, never, and no are a part of the negation dictionary. From the other person’s filter, the symptom will not be tagged if a sentence contains for example mother, brother etc. since it might relate to another person and not to the patient in question. The evaluation of the text mining was done using a confusion matrix of 200 randomly selected clinical notes. These were checked manually for incorrectly and correctly matched symptoms. Afterwards, the metrics sensitivity and specificity were calculated. The sensitivity is defined as

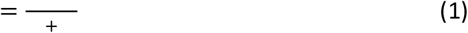

where TP are the true positives and FN the false negatives. The sensitivity describes the fraction of correctly matched symptoms opposed to all symptoms. Specificity is the fraction of correctly matched words that are not symptoms opposed to all the non-symptom words in the clinical notes and is defined by

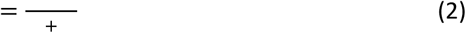

### Generating pancreas cancer trajectories

We calculated significant disease trajectories for the population of pancreatic cancer patients using the methodology from Jensen *et al*.*(A. B. Jensen et al. 2014)*. Here, the Relative Risk (RR) was calculated (**Eq. 3**) for the strength of an association between two diseases in the exposed group compared to a comparison group. The exposed group was matched with the same age and sex group as the comparison group and seasonal changes were accounted for by taking samples from the comparison group disease discharge to have the same week as the disease 1 (D1) discharge in the exposed group. The count for the exposed group is denoted C_exposed_ and the corresponding i’th comparison group as C_i_ where i ϵ {1,..,N} and N is the number of comparison groups.

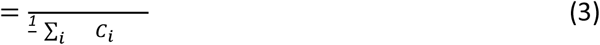

P-values for the RR were calculated using binomial tests, where the average probability of sampling a control patient with D2 was compared with C_exposed_. Afterwards, the diagnosis pairs (D1, D2) were tested for directionality with binomial tests to decide if the direction is significant for the significant disease pairs found. The patients having the direction (D1→ D2) or the other direction (D2→ D1) were counted. Patients having D1 and D2 at the same time were also counted and the total count constitutes N samples with 50 % probability of having either one of the directions. The p-values were afterwards Bonferroni corrected.

To find significant text mined symptoms, we sampled 10 control patients for every pancreatic cancer case patient. The 44,418 control patients were stratified based on sex, birth year and age at diagnosis. We ensured that the control patients were diagnosed with another disease at the same age as the cases and extracted their clinical notes five years previous to that diagnosis. This was done to make sure the notes for the control and the case group originated at a similar time period during their lifetime and up to a diagnosis.

For the construction of the symptom trajectories the RR was calculated using the counts for the co-occurrence of all possible symptom pair (See **Table 2**) and the *χ*^*2*^ was used to test the significance of the co-occurrence.

**Table 2.** Relative risk for symptoms pair.

| Relative Risk | $C_{\text{Exposed}}$ | $C_i$ |
| --- | --- | --- |
| $S_1 S_2$ | N patients in $C_{\text{Exposed}}$ with $S_1$ and $S_2$ | N patients in $C_i$ with $S_1$ and $S_2$ |
| Not $S_1 S_2$ | N patients in $C_{\text{Exposed}}$ without $S_1$ and $S_2$ | N patients in $C_i$ without $S_1$ and $S_2$ |

Subsequently, each symptom pair, with RR>1 and a significant p-value < 0.05, was tested for directionality and Bonferroni-corrected using the method from Jensen *et al*.*(A. B. Jensen et al. 2014)*. For the text mined symptoms, the day of admission was used as the time registered for the symptom, since capturing the symptom as early as possible is crucial for the purpose of this study.

## Data Availability

Permission to access the person-sensitive data used for this study can be obtained through the Danish Data Protection Agency, the Danish Health Authority, and the Danish Health regions (Capital Region and Region Zealand). Due to the sensitivity of these patient data, we have only provided diagnosis and co-occurrence information when grouped to at least five patients. All patient information published here is non-person-sensitive summary level data.

## Approvals

The work was approved as a registry study that does not require ethical permissions in Denmark as well as patient consent. Access to the data was approved by the Danish Data Protection Agency (ref: 514-0255/18-3000, 514-0254/18-3000, SUND-2016-50), the Danish Health Data Authority (ref: FSEID-00003724 and FSEID-00003092), and the Danish Patient Safety Authority (3-3013-1731/1/).

## Data availability

Permission to access the person-sensitive data used for this study can be obtained through the Danish Data Protection Agency, the Danish Health Authority, and the Danish Health regions (Capital Region and Region Zealand). As the raw electronic patient records and registry information are individual-level data they are person sensitive and cannot be made publicly available but only analyzed in closed, secure environments. In the paper we have only provided diagnosis and co-occurrence information when grouped to at least five patients. All patient information published here is non-person-sensitive summary level data and have been shared in the Supplementary materials.

## Code availability

The methodology and the data analysis have been carried out using Python software (version 3.8) and R version 3.6. The code for the text mining method is available at https://github.com/larsjuhljensen/tagger. The key algorithm for creation of disease trajectories have been described in details in the published studies Jensen *et al*.(Jensen et al. 2014) and *Siggaard et al*.(Siggaard et al. 2020).

## Acknowledgements

We would like to acknowledge funding from the Novo Nordisk Foundation (grant agreements NNF14CC0001 and NNF17OC0027594), the Braindrugs (R279-2018-1145) as well as the Danish Innovation Fund (5184-00102B).

## Author contributions

J.X.H. and SB conceptualized the study design and supervised the project. J.X.H. and S.I.N. ran the analyses together and wrote the manuscript draft together. T.S. helped run the text mining analysis. I.F.J. contributed in the decision making of the analysis and S.V. validated the text mining and translated the ICD-10 symptom chapter. J.S.J. and I.M.C. contributed to clinical expertise on pancreatic cancer oncology in Denmark. SB edited the draft version that was approved by all authors.

## Supplementary Tables

**Supplementary Table S1.** Complete names of ICD-10 codes that has been shortened for overview

| Abbreviation | Complete name |
| --- | --- |
| Nervous and musculoskeletal systems | Other and unspecified symptoms and signs involving the nervous and musculoskeletal systems |
| Abnormal findings on diagnostic images | Abnormal findings on diagnostic images of other body structures |
| Disorders of lipoprotein metabolism | Disorders of lipoprotein metabolism and other lipidaemias |
| Hypertension | Essential primary hypertension |
| Irregular menstruation | Excessive, frequent and irregular menstruation |
| Hearing loss | Other hearing loss |
| Digestive system and abdomen | Systems and signs involving digestive system and abdomen |
| Fever | Fever of other and unknown origin |
| Hyperalimentation | Other hyperalimentation |
| Pain | Pain, not elsewhere classified |
| Intestinal disorders | Other functional intestinal disorders |
| Oedema | Oedema, not elsewhere classified |
| Anaemia | Other anaemias |
| Jaundice | Unspecified jaundice |
| Paralytic ileus and intestinal obstruction | Paralytic ileus and intestinal obstruction without hernia |
| Haemorrhage | Haemorrhage, not elsewhere classified |
| Intestinal disorders | Other functional intestinal disorders |
| Abnormal blood pressure | Abnormal blood-pressure reading, without diagnosis |
| Soft tissue disorders | Other soft tissue disorders, not elsewhere classified |
| Elevation of levels of transaminase and LDH | Elevation of levels of transaminase and lactic acid dehydrogenase (LDH) |
| Food and fluid intake | Symptoms and signs concerning food and fluid intake |

**Supplementary Table S2.** Disease trajectories listing the number of patients following each trajectory and the median survival in days for each group.

| Trajectories | Number of patients | Median survival (days) |
| --- | --- | --- |
| C25_R50_R52 | 330 | 395 |
| E11_C25_R11 | 311 | 161 |
| E11_C25_R52 | 607 | 156 |
| E11_C25_R53 | 379 | 177 |
| E11_C25_R67 | 442 | 159,5 |
| E78_C25_R52 | 389 | 158,5 |
| H25_C25_R52 | 448 | 116 |
| H25_C25_R53 | 304 | 110 |
| H25_C25_R67 | 378 | 110,5 |
| H25_H26_C25 | 321 | 66 |
| H91_C25_R52 | 394 | 136 |
| H91_C25_R67 | 301 | 130 |
| I10_C25_K59 | 558 | 156 |
| I10_C25_R11 | 645 | 156 |
| I10_C25_R52 | 1073 | 156 |
| I10_C25_R53 | 737 | 161 |
| I10_C25_R63 | 484 | 160 |
| I10_C25_R67 | 831 | 146,5 |
| I20_C25_R52 | 441 | 161 |
| I20_C25_R67 | 312 | 140,5 |
| I20_E11_C25 | 302 | 84 |
| I20_E78_C25 | 413 | 91,5 |
| I20_I25_C25 | 888 | 73 |
| I20_R10_C25 | 301 | 92 |
| I21_I20_C25 | 599 | 73 |
| I21_I20_I25 | 373 | 78 |
| I21_I25_C25 | 909 | 65 |
| I21_I50_C25 | 363 | 48 |
| I25_C25_R52 | 372 | 133 |
| K80_C25_R52 | 458 | 187 |
| K80_C25_R67 | 322 | 190 |
| K85_K86_C25 | 340 | 96,5 |
| M17_C25_R52 | 348 | 189 |
| M17_I10_C25 | 332 | 68 |
| M51_C25_R52 | 320 | 182 |
| M51_I10_C25 | 301 | 91,5 |
| M54_C25_R52 | 353 | 170 |
| M79_C25_R52 | 306 | 187 |
| N92_C25_R52 | 394 | 185 |
| N92_R10_C25 | 304 | 102 |
| R10_C25_K59 | 530 | 156 |
| R10_C25_R11 | 600 | 176 |
| R10_C25_R52 | 1029 | 174 |
| R10_C25_R53 | 656 | 177 |
| R10_C25_R63 | 438 | 196 |
| R10_C25_R67 | 738 | 169 |

**Supplementary Table S3.**
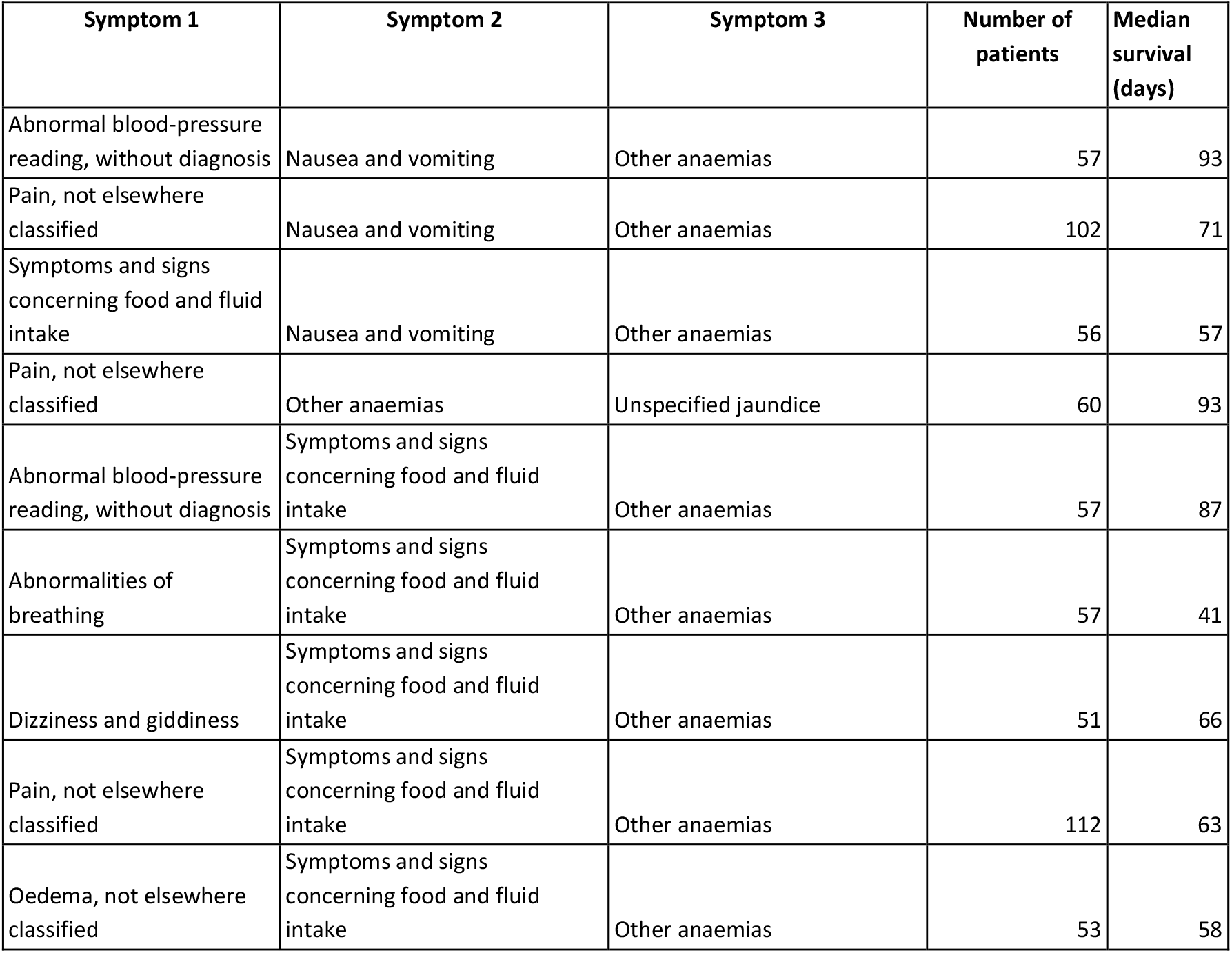
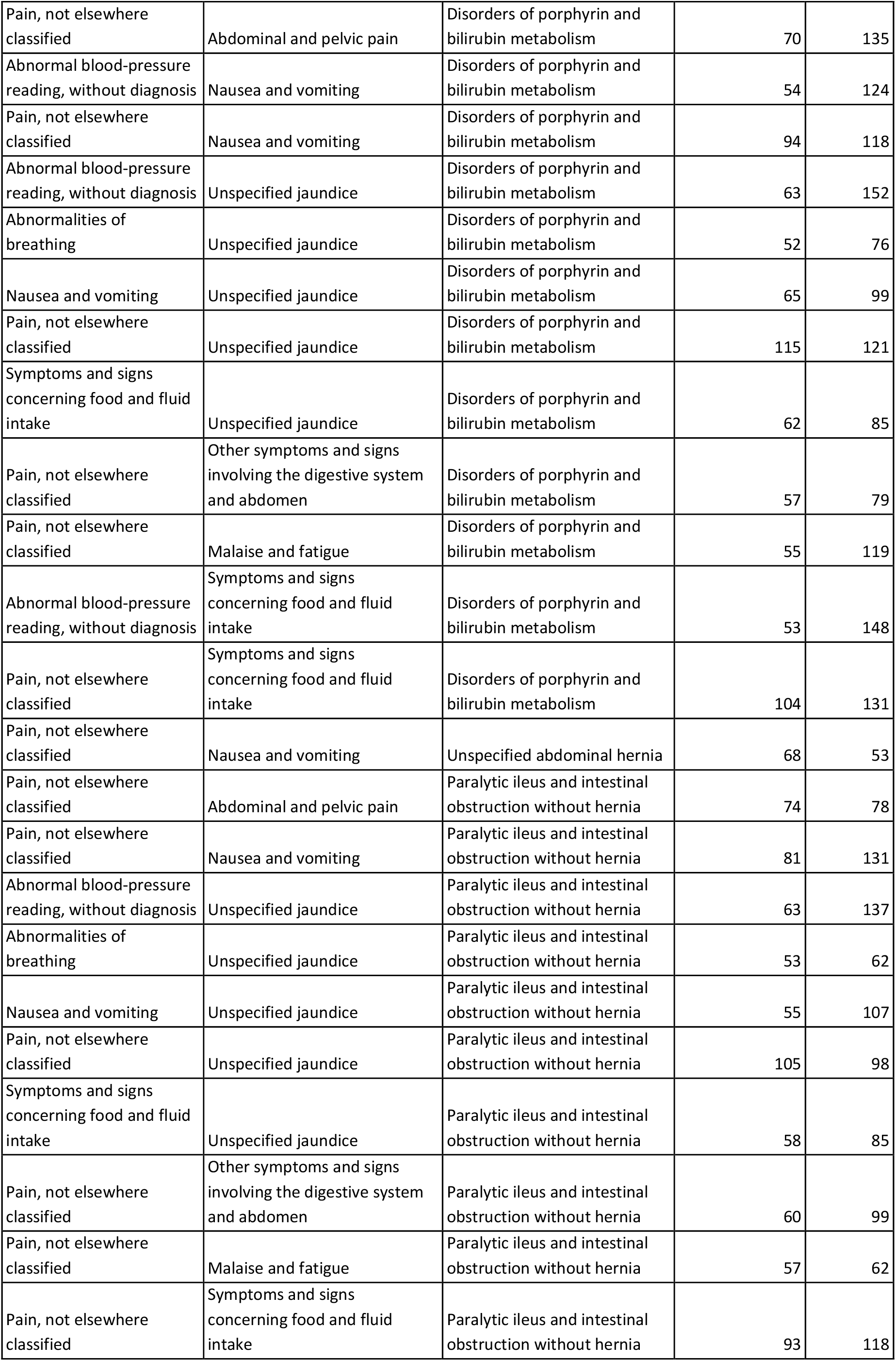
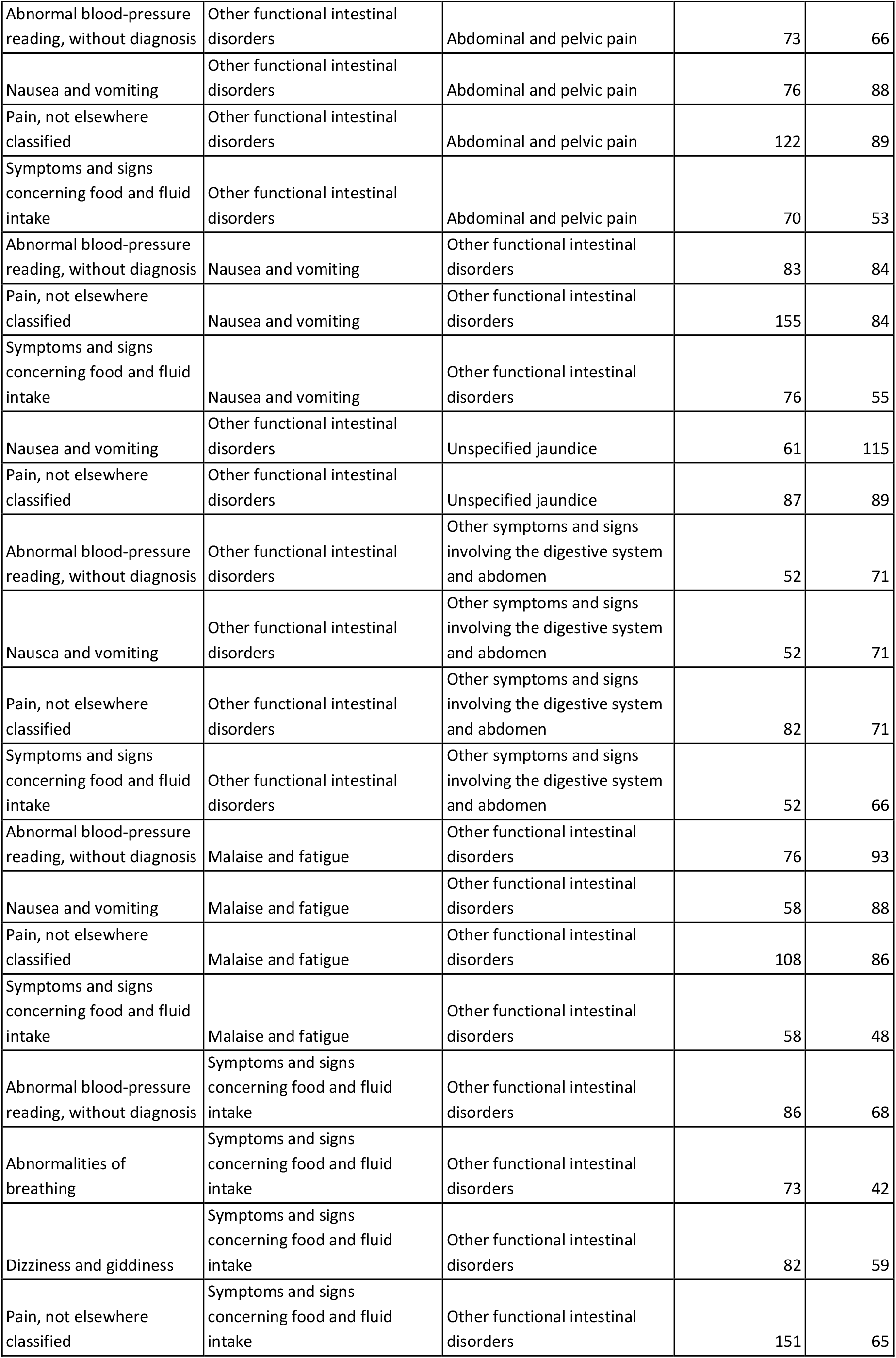
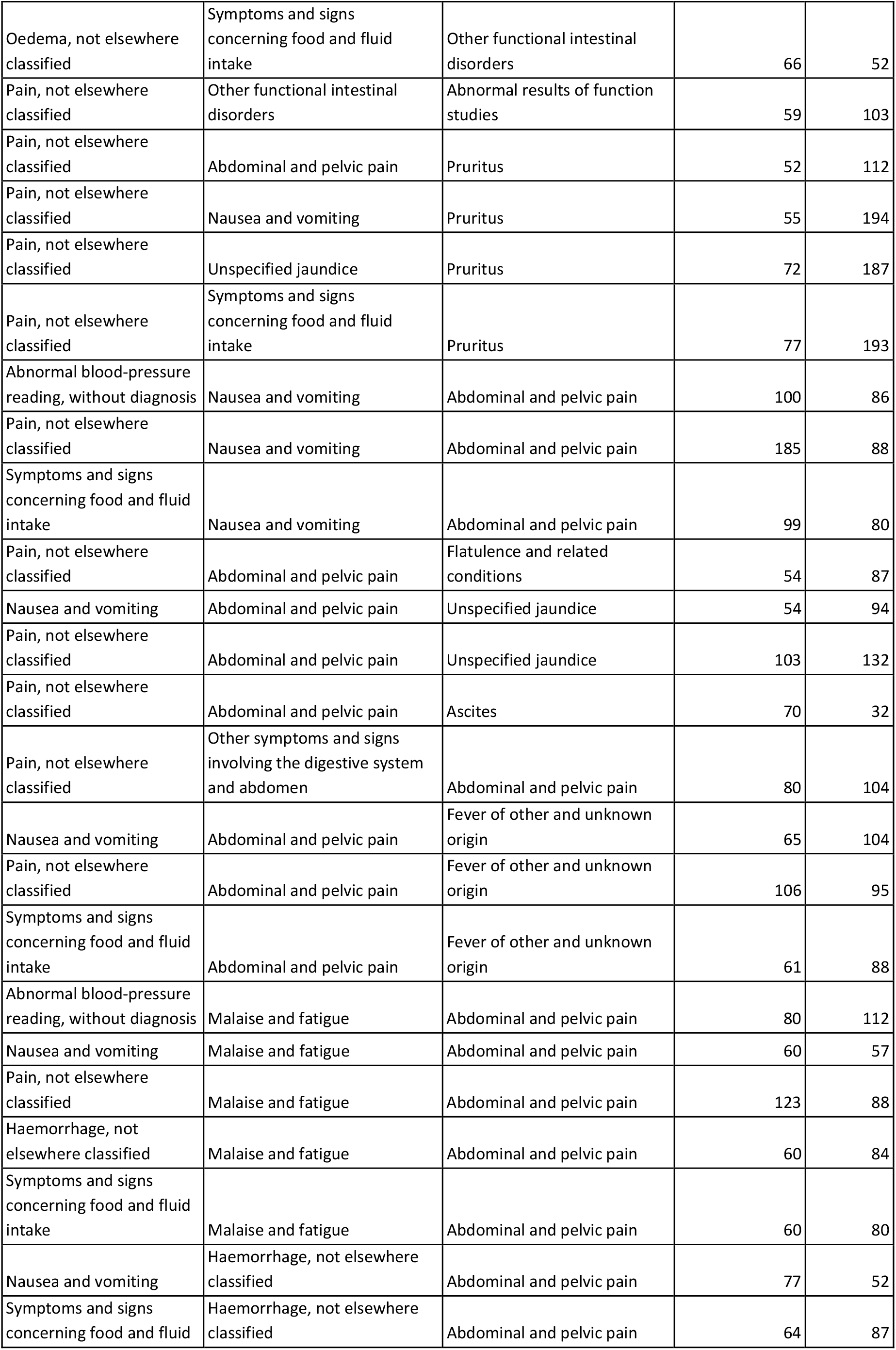
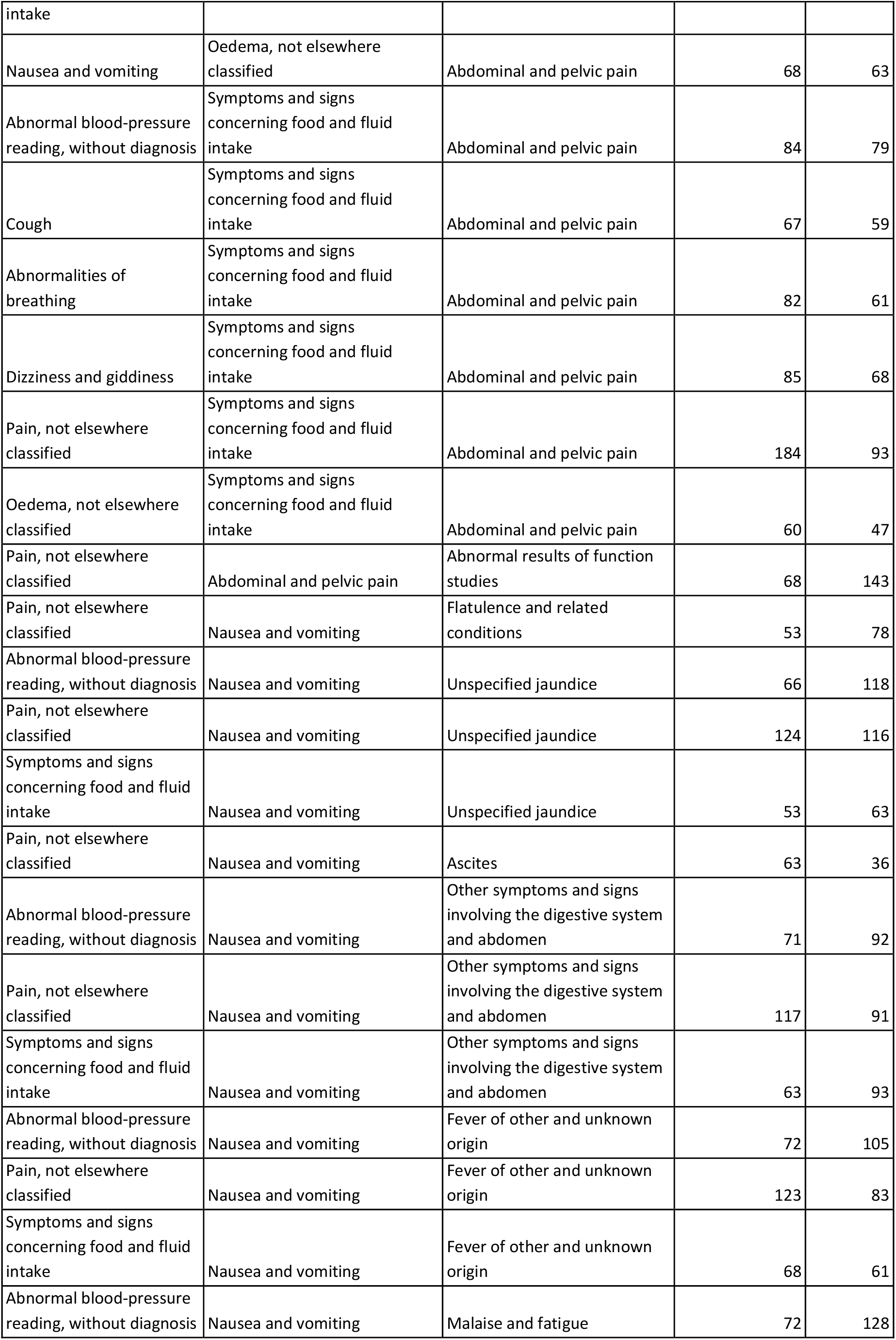
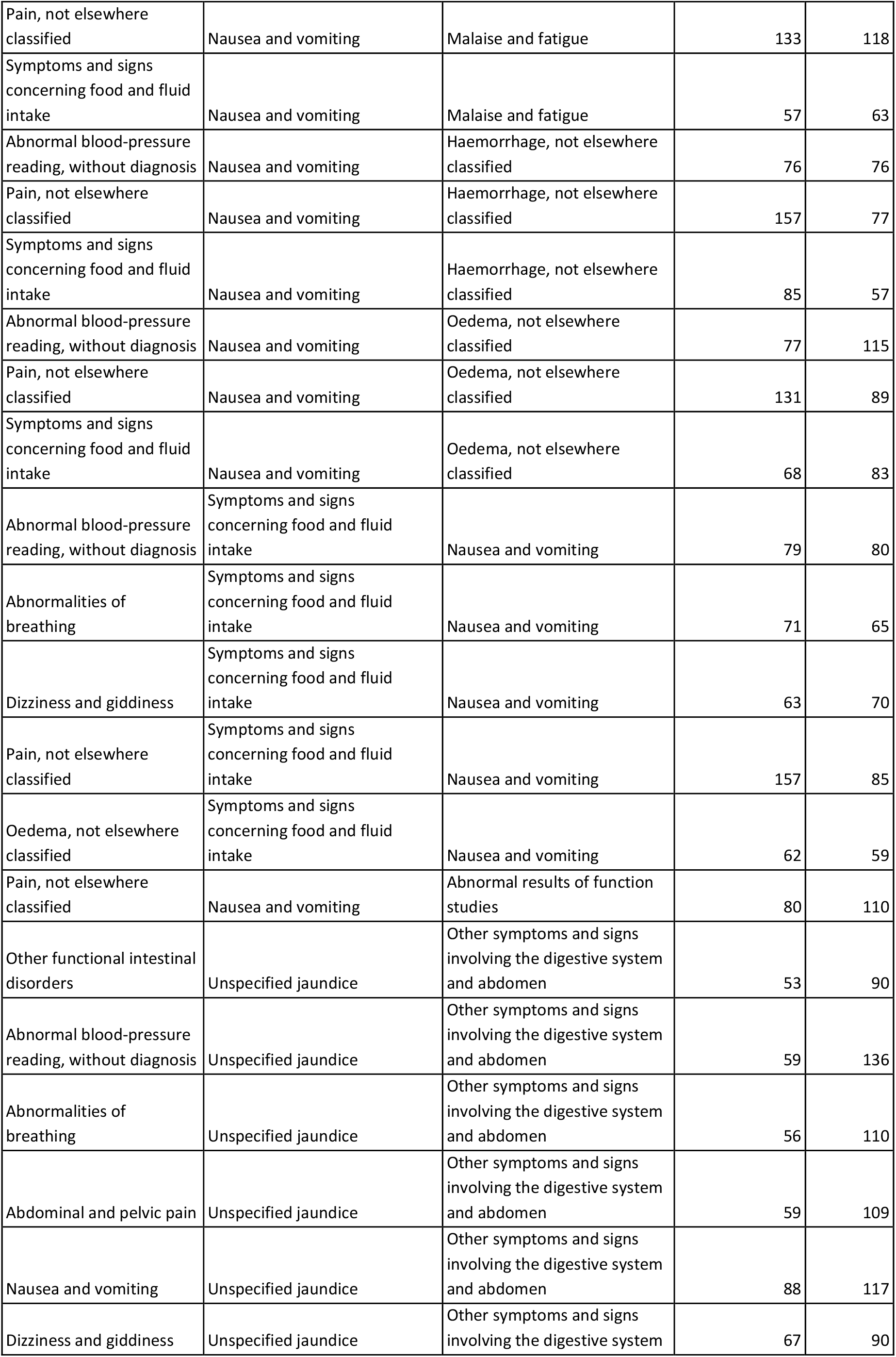
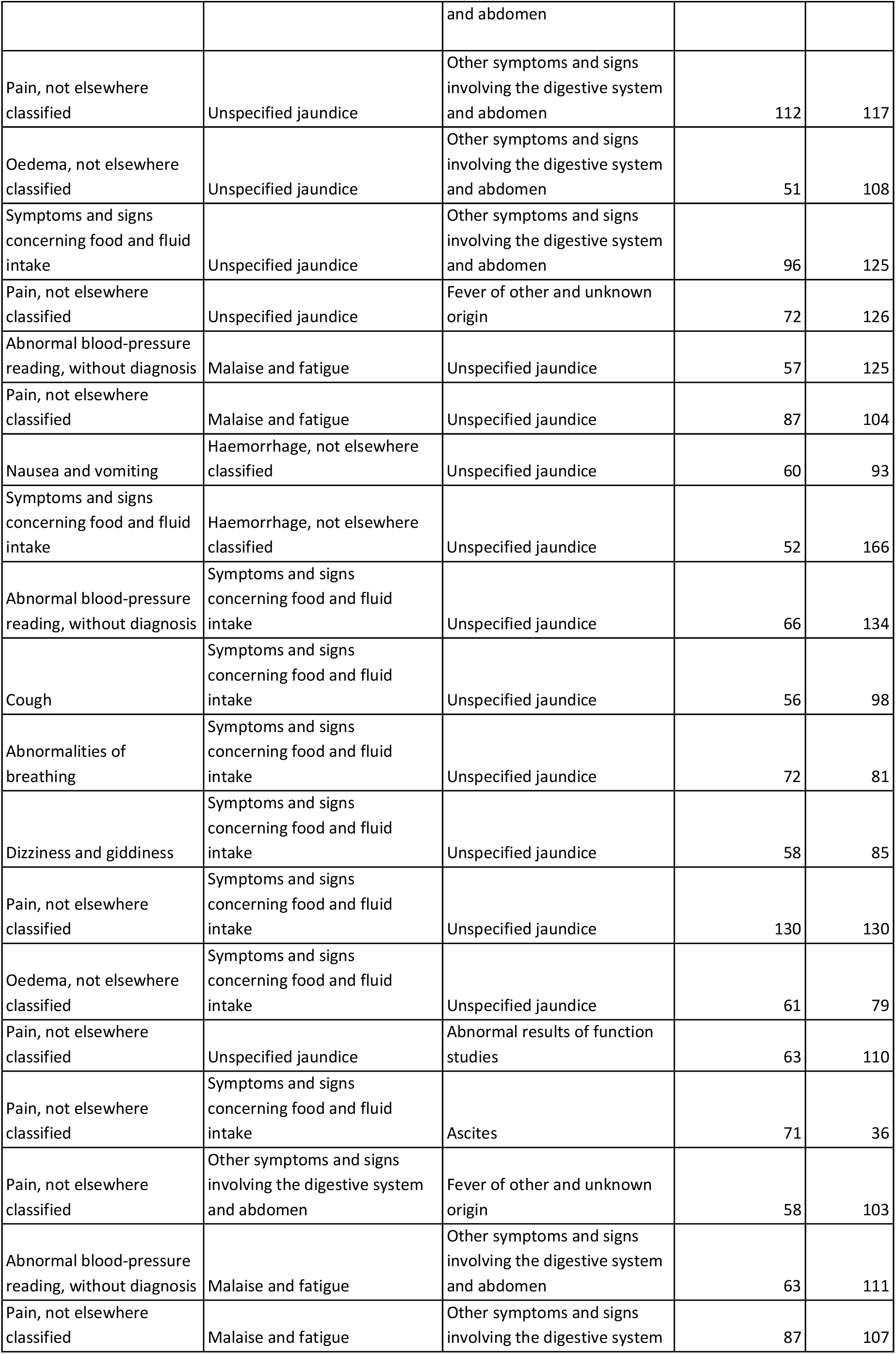
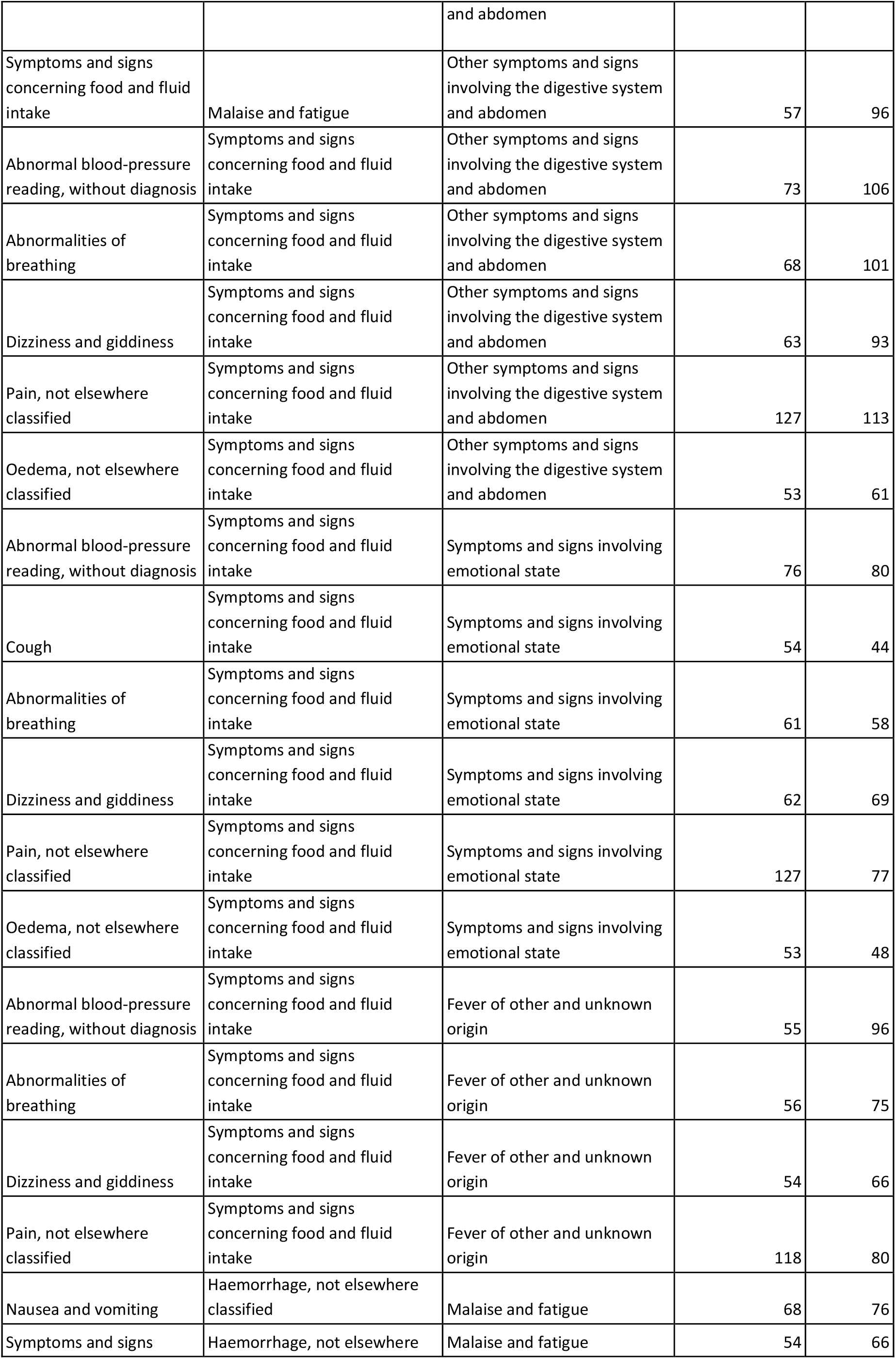
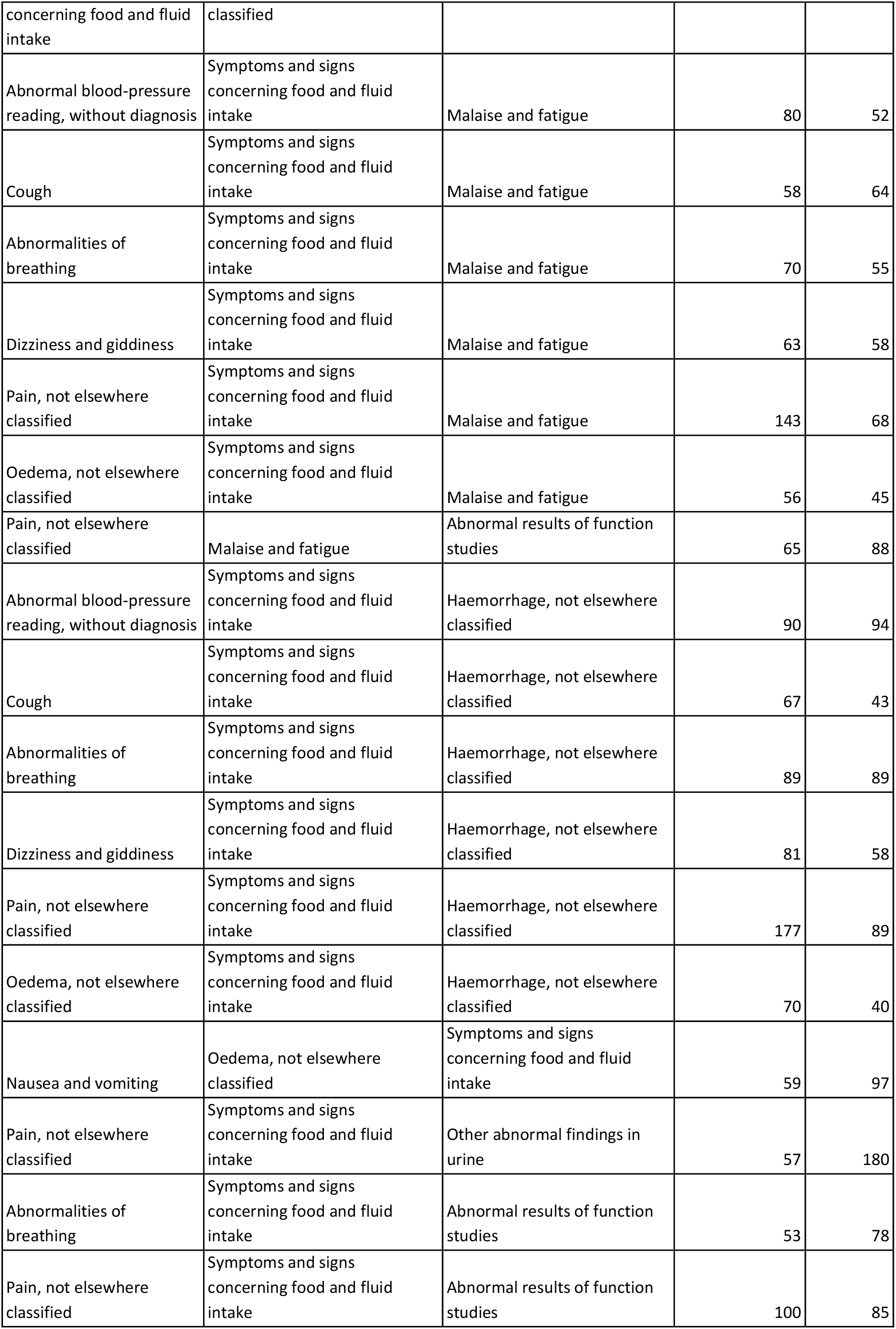
Symptom trajectories listing the number of patients following each trajectory and the median survival in days for each group. The table lists all symptom trajectories found with a minimum of 50 patients.

**Supplementary Table S4.** The 20 most frequent symptoms found by text mining clinical notes. Significance is tested by a *χ*^*2*^ test statistics.

| Symptom | Number of patients | P-value |
| --- | --- | --- |
| Jaundice | 1184 | $0.00 \cdot 10^{00}$ |
| Anorexia | 1374 | $0.00 \cdot 10^{00}$ |
| Digestive system and abdomen | 1033 | $1.44 \cdot 10^{-146}$ |
| Abnormal weight loss | 2111 | $0.00 \cdot 10^{00}$ |
| Food and fluid intake | 2468 | $0.00 \cdot 10^{00}$ |
| Abdominal and pelvic pain | 1400 | $1.54 \cdot 10^{-250}$ |
| Nausea | 1551 | $4.32 \cdot 10^{-91}$ |
| Nausea and vomiting | 1858 | $9.21 \cdot 10^{-93}$ |
| Vomiting | 1085 | $1.06 \cdot 10^{-44}$ |
| Malaise and fatigue | 1171 | $2.73 \cdot 10^{-29}$ |
| Fever | 796 | $1.45 \cdot 10^{-20}$ |
| Intestinal disorders | 1045 | $1.63 \cdot 10^{-24}$ |
| Functional diarrhoea | 685 | $2.47 \cdot 10^{-16}$ |
| Other anaemias | 658 | $4.44 \cdot 10^{-09}$ |
| Haemorrhage | 1520 | $4.80 \cdot 10^{-08}$ |
| Pain | 2982 | $2.21 \cdot 10^{-15}$ |
| Abnormal blood-pressure | 1677 | $7.28 \cdot 10^{-03}$ |
| Abnormalities of breathing | 1221 | $1.90 \cdot 10^{-04}$ |
| Headache | 619 | $2.12 \cdot 10^{-06}$ |
| Abnormalities of heart beat | 641 | $1.33 \cdot 10^{-07}$ |

**Supplementary Table S5.**
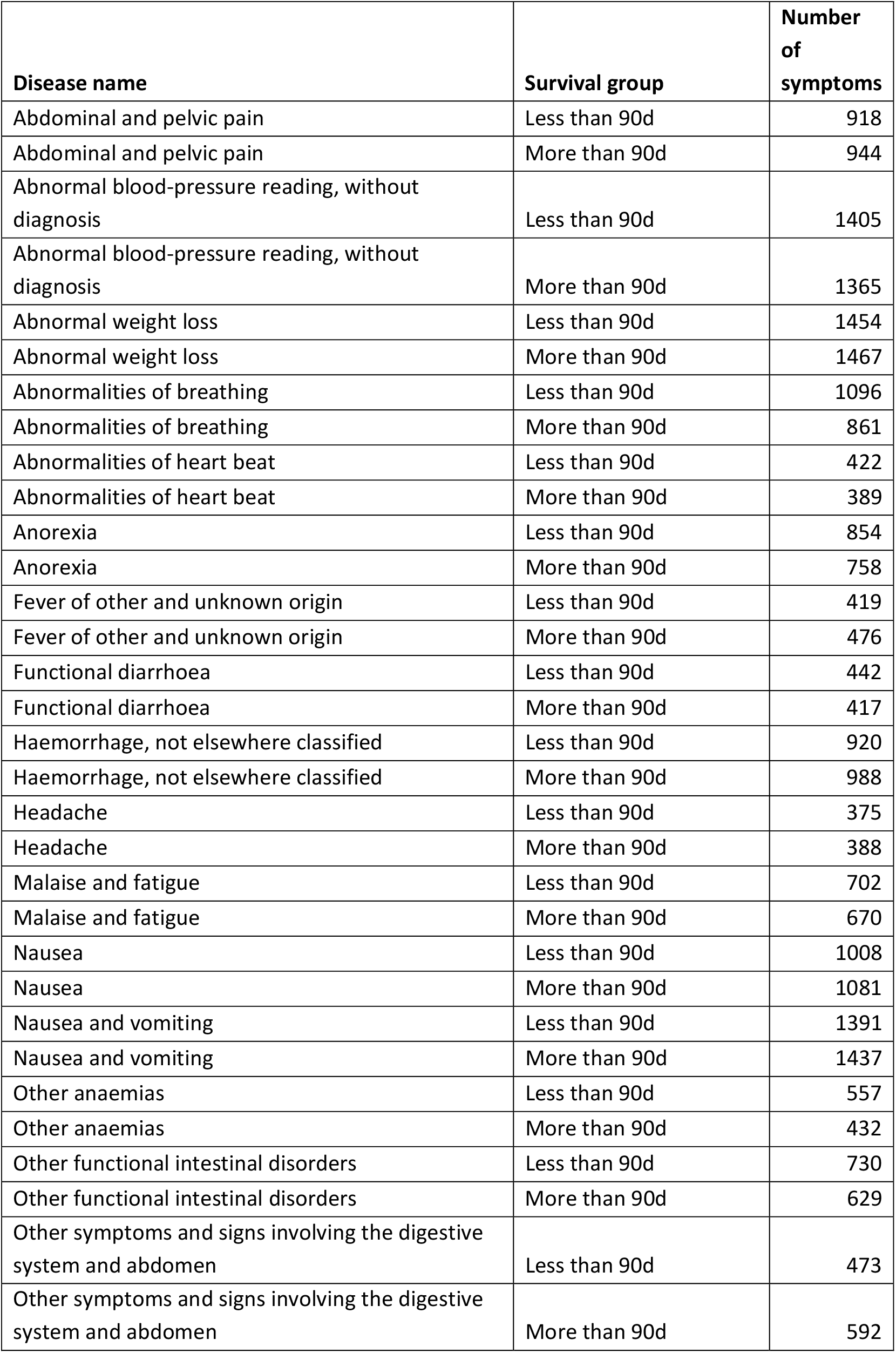
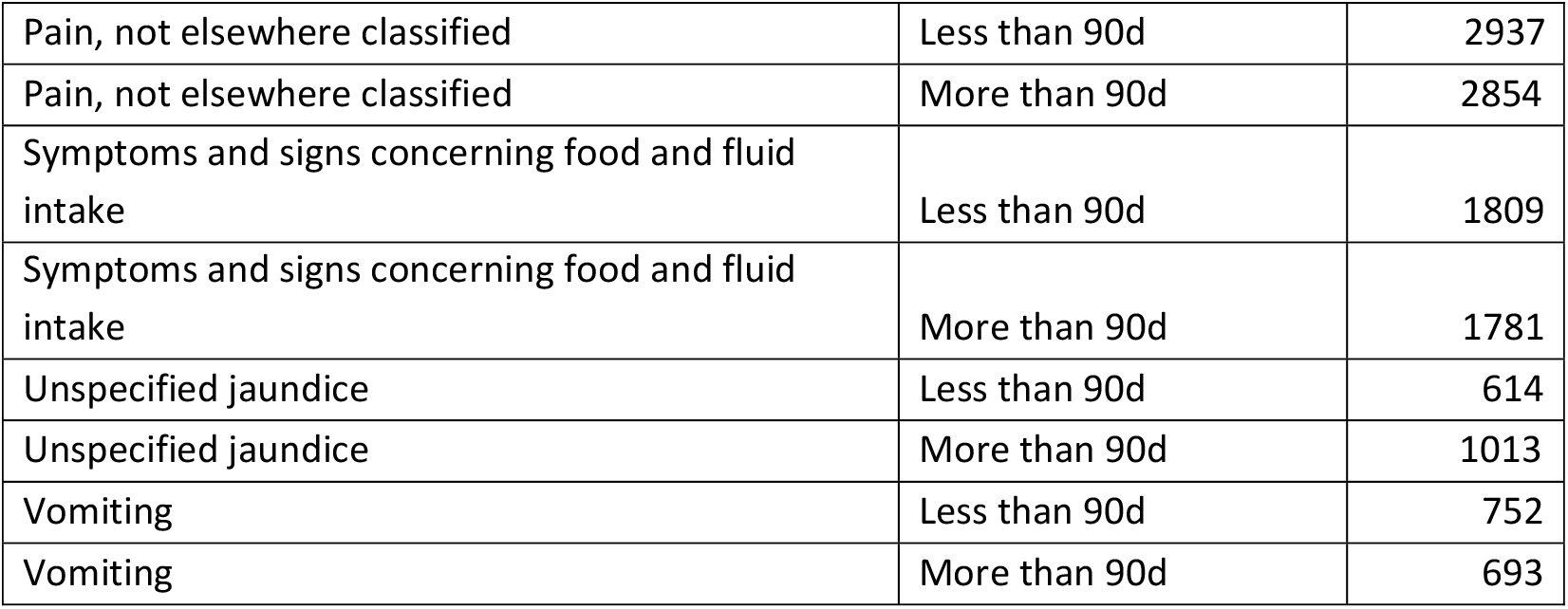

## Supplementary Figure

**Figure S1.**
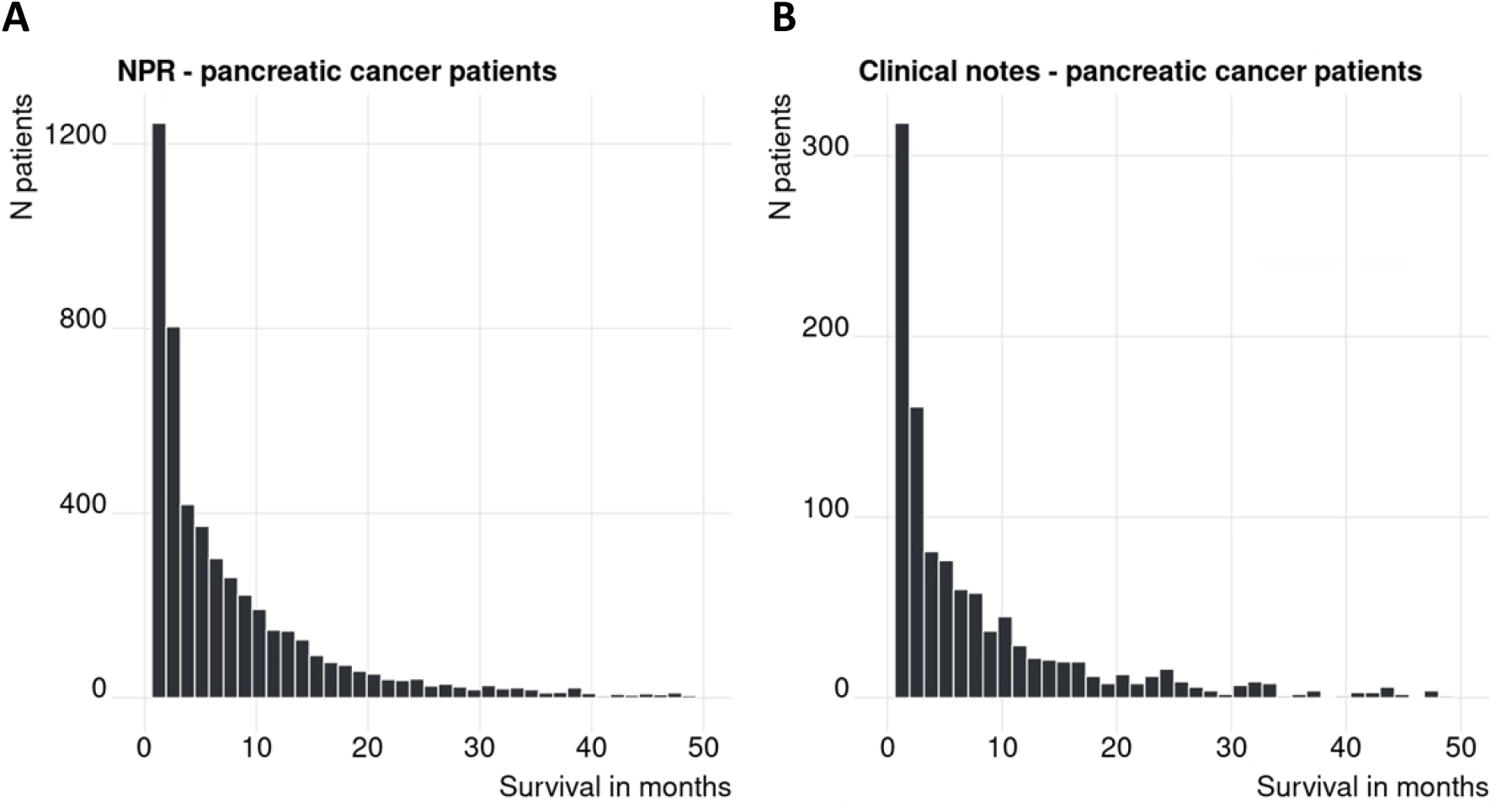
(A) Survival in months for patients in the registry disease trajectories. (B) Survival in months for patients in the text mining symptom trajectories. The pancreatic cancer patient cohort from is a subset of the pancreatic cancer patient cohort in A.

